# Low uptake of COVID-19 lateral flow testing among university students: a mixed methods evaluation

**DOI:** 10.1101/2021.07.20.21260836

**Authors:** C.E. French, S. Denford, E. Brooks-Pollock, H. Wehling, M. Hickman

## Abstract

**Objective:** To evaluate COVID-19 lateral flow testing (LFT) among asymptomatic university students.

**Study design:** Mixed methods evaluation of LFT among University of Bristol students.

**Methods:** i) An analysis of testing uptake and exploration of demographic variations in uptake using logistic regression; ii) an online student survey about views on university testing; and iii) qualitative interviews to explore participants’ experiences of testing and subsequent behaviour, analysed using a thematic approach.

**Results:** 12,391 LFTs were conducted on 8025/36,054 (22.3%) students. Only one in 10 students had the recommended two tests. There were striking demographic disparities in uptake with those from ethnic minority groups having lower uptake (e.g. 3% of Chinese students were tested vs. 30.7% of White students), and variations by level and year of study (ranging from 5.3% to 33.7%), place of residence (29.0% to 35.6%) and faculty (15.2% to 32.8%). Differences persisted in multivariable analyses.

A total of 436 students completed the online survey, and twenty in-depth interviews were conducted. Barriers to engagement with testing included a lack of awareness, knowledge and understanding, and concerns about the accuracy and safety. Students understood limitations of LFTs but requested further information about test accuracy. Tests were used to inform behavioural decisions, often in combination with other information, such as the potential for exposure to the virus and perceptions of vulnerability.

**Conclusions:** The low uptake of testing brings into question the role of mass LFT in university settings. Innovative strategies may be needed to increase LFT uptake among students.

## INTRODUCTION

Lateral flow testing (LFT) of asymptomatic people remains an integral part of the UK’s COVID-19 response. Since 9^th^ April 2021 everyone in England has been eligible to take a LFT twice weekly ^1-4^. There is an ongoing and polarised debate around mass testing to detect asymptomatic infections using this technology. Since approximately one third of people infected with SARS-CoV-2 have no symptoms, it is argued that identifying infections among this group so that they can isolate and their contacts be traced is key to controlling the pandemic ^3, 4^. Although this policy was well received by some ^5-7^ others have raised concerns, particularly around test accuracy and the potential consequences of inaccurate results ^8-11^. While the accuracy of LFT is important, much less attention has been paid to levels of uptake of testing, which could pose a major barrier for the use and effectiveness of asymptomatic testing.

In Autumn 2020, COVID cases were high among university students in the UK ^12^. In November 2020 the government recommended LFT for university students, recommending that all students should have two negative tests before travelling home for the winter break ^1, 13^. Evaluation of this testing strategy, including equity in testing uptake is crucial if testing continues to be used to control the pandemic in the future.

University populations offer a unique opportunity to quantify testing uptake in a well-defined group of individuals. Our study aims to i) assess uptake of LFT among University of Bristol students, including demographic variations; ii) explore the acceptability and feasibility of asymptomatic testing and iii) to explore the barriers and facilitators to uptake and effective implementation of testing.

## METHODS

We conducted a mixed methods evaluation of LFT among University of Bristol students who did not have COVID-19 symptoms; comprising a quantitative analysis of testing uptake data, a student survey and qualitative interviews.

### Testing uptake

We analysed data on the uptake of LFT from 30^th^ November to 18^th^ December 2020. Students pre-booked their tests online. On arrival at testing venues they were asked to swipe their university identity card. A list of all students enrolled at the university, held by student records, was matched with the date of any tests undertaken, as collected via card swipes at testing venues using student ID number. Information on location of students during the study period was not available. However, a sensitivity analysis was conducted by excluding students who were either enrolled on a distance learning course or completed a ‘location of study’ form indicating that they were likely not going to be on campus. The total number of positive results was recorded at testing sites but was not documented for individual students. Univariable and multivariable analyses were conducted using logistic regression to explore demographic factors associated with being tested. All explanatory variables were included in the multivariable model *a priori*. Analyses were conducted in STATA 16.1 (StataCorp LLC, College Station, TX).

### Survey

Participants were invited to complete a confidential online survey about their views of university testing (Supplement 1). A link to the survey was shared by the university communications team via social media (Facebook, Twitter and Instagram) and via the student newsletter. Informed consent was obtained.

Frequencies and descriptive statistics are presented for closed survey questions. Free text answers were used to offer further insight into answers given to closed survey questions. We identified key barriers to engagement with testing using qualitative content analysis in three stages ^14-16^ – survey responses were coded independently by two authors, codes were then categorised into a list of barriers and facilitators, and data assigned to each category.

### Interviews

Volunteers who took part in the survey and provided consent to be contacted by the research team were invited to take part in an online interview. Participants were >18 years and a registered student at the university. We purposely sampled for diversity in key factors, including ethnicity, living arrangements, enrolled course, and whether or not they had taken a test at the university. Sample size was informed by the concept of ‘information power’ ^17^, with continuous assessment of the data in relation to study objectives.

Potential participants were provided with a study information sheet and given an opportunity to ask questions, informed of the voluntary nature of the study, and assured of the confidentiality of their data. As all interviews were conducted via the telephone or online, and audio recorded verbal consent was obtained.

The semi-structured topic guide (Supplement 2) aimed to explore participants’ views about testing, understanding and interpretation of test results, and impact on behaviour.

Data from interviews were analysed using a thematic approach ^18, 19^. Two researchers independently read and assigned codes to transcripts. Possible themes were identified and refined. Charts were developed for each theme, and relevant text from transcripts were copied verbatim. Charts were then used to compare data within and between individuals.

## RESULTS

### Quantitative analysis

#### Testing uptake

A total of 12,391 LFD tests were conducted on 8025 (22.3%) of the 36,054 students enrolled at the university. Of those tested, 3921 (48.9%) had one test, 3880 (48.3%) had the recommended two tests, 189 (2.4%) had three tests and 35 (0.4%) had four to six tests. There were 13 positive results.

#### Demographic variations in testing uptake (Tables 1 and 2)

Although the absolute percentage of students taking up testing was similar across genders (21.9% for men and 22.5% for women), women were more likely to be tested than men (adjusted odds ratio [aOR]: 1.18, 95% confidence interval [CI]: 1.11-1.25). There were striking variations in uptake by ethnic group. Uptake was highest in ethnically White students, with 30.7% taking at least one test. Uptake was lower among all other groups - it was lowest among students belonging to the Chinese ethnic group (3%, aOR: 0.17, 95% CI: 0.14-0.20), followed by the Black African, Black Caribbean and Black other group (12.3%, aOR: 0.34, 95% CI: 0.28-0.42). It was also low among the Indian, Pakistani and Bangladeshi group (17.5%, aOR: 0.53, 95% CI: 0.47-0.61).

**Table 1.** Demographic characteristics of students according to uptake of testing (n=36,054)

| Characteristic | Not tested |  | Tested |  | Total |
| --- | --- | --- | --- | --- | --- |
|  | n | % | n | % | n |
| **Gender** |  |  |  |  |  |
| Male | 12,430 | 78.1 | 3,489 | 21.9 | 15,919 |
| Female | 15,557 | 77.5 | 4,526 | 22.5 | 20,083 |
| Other | 40 | 80.0 | 10 | 20.0 | 50 |
| **Ethnic group** |  |  |  |  |  |
| White | 14,675 | 69.3 | 6,508 | 30.7 | 21,183 |
| Indian, Pakistani, Bangladeshi | 1,423 | 82.5 | 301 | 17.5 | 1,724 |
| Black African, Black Caribbean, Black other | 742 | 87.7 | 104 | 12.3 | 846 |
| Chinese | 5,543 | 97.0 | 172 | 3.0 | 5,715 |
| Mixed | 1,220 | 72.2 | 470 | 27.8 | 1,690 |
| Other | 1,464 | 86.9 | 220 | 13.1 | 1,684 |
| Not reported | 2,962 | 92.2 | 250 | 7.8 | 3,212 |
| **Level of study** |  |  |  |  |  |
| Undergraduate | 15,700 | 69.3 | 6,960 | 30.7 | 22,660 |
| Postgraduate - Research | 3,645 | 86.4 | 575 | 13.6 | 4,220 |
| Postgraduate - Taught | 8,684 | 94.7 | 490 | 5.3 | 9,174 |
| **Year of study\*** |  |  |  |  |  |
| Year 1** | 5,898 | 72.6 | 2,225 | 27.4 | 8,123 |
| Year 2 | 4,384 | 68.4 | 2,025 | 31.6 | 6,409 |
| Year 3 | 3,873 | 66.8 | 1,926 | 33.2 | 5,799 |
| Year 4+ | 1,545 | 66.3 | 784 | 33.7 | 2,329 |
| **Place of residence\*** |  |  |  |  |  |
| In halls | 3,779 | 64.4 | 2,093 | 35.6 | 5,872 |
| Not in halls | 11,921 | 71.0 | 4,867 | 29.0 | 16,788 |
| **Faculty** |  |  |  |  |  |
| Faculty of Science | 2,945 | 67.2 | 1,438 | 32.8 | 4,383 |
| Faculty of Arts | 4,833 | 74.1 | 1,694 | 26.0 | 6,527 |
| Faculty of Engineering | 4,267 | 81.6 | 960 | 18.4 | 5,227 |
| Faculty of Health Sciences | 3,232 | 75.1 | 1,072 | 24.9 | 4,304 |
| Faculty of Life Sciences | 2,712 | 71.8 | 1,065 | 28.2 | 3,777 |
| Faculty of Social Science and Law | 10,039 | 84.8 | 1,796 | 15.2 | 11,835 |

\*Restricted to undergraduate students only
\*\*Includes 153 pre-sessional students

**Table 2.** Univariable and multivariable logistic regression analyses of demographic characteristics associated with testing uptake.

| Characteristic | Univariable analysis |  |  | Multivariable analysis (n=36,051) |  |  |
| --- | --- | --- | --- | --- | --- | --- |
|  | Odds ratio* | 95% CI | p value | Adjusted odds ratio* | 95% CI | p value |
| **Gender** |  |  |  |  |  |  |
| Male | Reference |  |  | Reference |  |  |
| Female | 1.04 | 0.99-1.09 | 0.161 | 1.18 | 1.11-1.25 | <0.001 |
| Other | 0.89 | 0.44-1.78 | 0.744 | 1.42 | 0.67-3.02 | 0.360 |
| **Ethnic group** |  |  |  |  |  |  |
| White | Reference |  |  | Reference |  |  |
| Indian, Pakistani, Bangladeshi | 0.48 | 0.42-0.54 | <0.001 | 0.53 | 0.47-0.61 | <0.001 |
| Black African, Black Caribbean, Black other | 0.32 | 0.26-0.39 | <0.001 | 0.34 | 0.28-0.42 | <0.001 |
| Chinese | 0.07 | 0.06-0.08 | <0.001 | 0.17 | 0.14-0.20 | <0.001 |
| Mixed | 0.87 | 0.78-0.97 | 0.012 | 0.84 | 0.75-0.95 | 0.004 |
| Other | 0.34 | 0.29-0.39 | <0.001 | 0.44 | 0.38-0.51 | <0.001 |
| Not reported | 0.19 | 0.17-0.22 | <0.001 | 0.20 | 0.17-0.22 | <0.001 |
| **Student group** |  |  |  |  |  |  |
| Undergraduate - Year 1** - In halls | Reference |  |  | Reference |  |  |
| Undergraduate - Year 1** - Not in halls | 0.13 | 0.11-0.15 | <0.001 | 0.20 | 0.17-0.24 | <0.001 |
| Undergraduate - Year 2 | 0.82 | 0.76-0.88 | <0.001 | 0.85 | 0.79-0.92 | <0.001 |
| Undergraduate - Year 3 | 0.88 | 0.82-0.95 | 0.001 | 0.88 | 0.81-0.95 | 0.001 |
| Undergraduate - Year 4+ | 0.90 | 0.81-1.00 | 0.042 | 0.85 | 0.76-0.95 | 0.004 |
| Postgraduate - Research | 0.28 | 0.25-0.31 | <0.001 | 0.28 | 0.25-0.31 | <0.001 |
| Postgraduate - Taught | 0.10 | 0.09-0.11 | <0.001 | 0.15 | 0.14-0.17 | <0.001 |
| **Faculty** |  |  |  |  |  |  |
| Faculty of Science | Reference |  |  | Reference |  |  |
| Faculty of Arts | 0.72 | 0.66-0.78 | <0.001 | 0.64 | 0.59-0.70 | <0.001 |
| Faculty of Engineering | 0.46 | 0.42-0.51 | <0.001 | 0.70 | 0.63-0.77 | <0.001 |
| Faculty of Health Sciences | 0.68 | 0.62-0.75 | <0.001 | 0.67 | 0.61-0.75 | <0.001 |
| Faculty of Life Sciences | 0.80 | 0.73-0.88 | <0.001 | 0.75 | 0.68-0.83 | <0.001 |
| Faculty of Social Sciences and Law | 0.37 | 0.34-0.40 | <0.001 | 0.63 | 0.58-0.69 | <0.001 |

\*An odds ratio of less than one indicates lower uptake of testing as compared with the reference group
\*\*Includes 153 pre-sessional students

When compared with Year 1 undergraduate students living in halls of residence, Year 1 undergraduate students not living in halls were less likely to be tested (aOR: 0.20, 95% CI:0.17-0.24), as were postgraduate students, particularly postgraduate taught students (aOR: 0.15, 95% CI:0.14-0.17). Testing uptake also varied by faculty. Compared with students in the Faculty of Science, uptake was lower among those in all other faculties. It was lowest in the Faculty of Social Sciences and Law and the Faculty of Arts.

A sensitivity multivariable analysis excluding students who were likely not to have been on campus during the testing period (n=4,907, 13.6% of all students) did not alter the observed patterns in testing uptake. Odds ratios changed a little (all <10%) and were within the confidence intervals reported in Table 2.

### Survey

A total of 436 students completed the survey, of which 328 (75%) had taken part in testing and 108 (25%) had not (Supplement 3).

#### Attitudes towards testing

Among students who engaged in the university testing service and those who did not, the majority described their views of getting regular tests as either somewhat positive (31% and 31% respectively) or very positive (51% v 31%). Few participants described their views of testing as somewhat negative or very negative (18% of those who did not participate in testing vs. 5% of those who did: Table 3).

**Table 3.** Responses to survey questions.

|  | **Participated in testing N**<br>**= 328** | **Did not participate in testing**<br>**N = 108** |
| --- | --- | --- |
| **Views on getting tested regularly** |  |  |
| Very negative | 2 (1%)* | 5 (5%)* |
| Somewhat negative | 14 (4%)* | 14 (13%)* |
| Neither positive or negative | 31 (9%) | 16 (14%) |
| Somewhat positive | 103 (31%) | 39 (31%) |
| Very positive | 169 (51%)* | 33 (31%)* |
| **Interpretation of negative test results** |  |  |
| The person is definitely infectious | 6 (2%) | 1 (1%) |
| The person is probably infectious | 11 (3%) | 3 (3%) |
| The person is probably not infectious | 277 (84%) | 81 (75%) |
| The person is definitely not infectious | 21 (6%)* | 13 (12%)* |
| Don't know | 13 (4%) | 10 (9%) |
| **Close contact following test** |  |  |
| Much more contact | 12 (4%) | NA |
| Slightly more contact | 49 (15%) | NA |
| About the same | 180 (55%) | NA |
| Slightly less | 22 (7%) | NA |
| Much less | 35 (10%) | NA |
| Missing | 30 | NA |
| **Adherence to social distancing recommendations** |  |  |
| All of the time | 139 (42%) | 41 (38%) |
| Most of the time | 156 (48%) | 47 (43%) |
| Some of the time | 19 (6%) | 7 (6%) |
| Not at all | 1 (0%) | 5 (5%) |
| Missing | 13 (4%) | 8 (7%) |

\*Denotes that group proportions differ significantly at the 0.05 level

#### Interpretation of test results

Most students understood that a negative test result meant that the person is probably not infectious (84% of those who had a test versus 75% of those who did not – Table 3). Only a minority of students in both groups thought a negative test means the person is definitely not infectious (6% of those engaging in testing v 12% of those who did not) or that they did not know (4% of those engaging in testing v 9% of those who did not).

#### Behaviour

Approximately half of students engaging in testing reported that the level of contact with others had not changed in the seven days following the testing period (55%). 19% of students reported that close contact increased, and 17% reported that close contact had decreased following tests (Table 3). Self-reported adherence to the guidance was similar between the groups, with 90% of those engaging in testing and 81% of those not engaging in testing reporting that they had been adherent to the guidance all or most of the time (Table 3).

#### Barriers

A total of 108 comments were coded and used to identify barriers to engagement in testing (Table 4). Barriers were categorised as 1) Perceived lack of need or demand 2) Problems accessing the service 3) Safety concerns 4) Knowledge and understanding and 5) Lack of support for self-isolation.

**Table 4.** Coded survey responses relating to barriers and facilitators to testing.

| Theme | Description | Example quote | Count |
| --- | --- | --- | --- |
| **Perceived lack of need / demand** |  |  |  |
| Lack of exposure/ self-isolating | Includes comments about not requiring tests due to not being exposed to the virus (e.g., as a result of students self-isolating) | *"I had already been isolating (by choice) for two weeks, so that I was able to go home"* | 6 |
| Lack of travel plans | Includes comments by participants who are not intending to leave Bristol | *"As I had no plans to go home over Christmas I didn't go for a test"* | 11 |
| (Low) priority | Captures comments by participants who do not think COVID is a threat | *Completely unnecessary, cancer has a higher chance of death but I don't get tested for cancer* | 1 |
| Students not in Bristol | Many students were not in Bristol at the time of testing | *I had already returned home for lockdown before tests were available* | 13 |
| Previously tested positive | Comments about tests not being necessary due to having previously tested positive | *"I have already had the virus so would not be expected to contract it again"* | 9 |
| **Accessing the service** |  |  |  |
| Location | Includes comments about testing sites being inaccessible to those who live off campus, are based at a different campus (e.g., Langford) and/or who are new to the University and not familiar with the layout. | *"Test site are too far away for many students in private housing"* | 12 |
| Timing of testing | Includes comments relating to a too narrow testing window for some students – in particular international students, those on placement, and/or those with jobs were not able to travel within the window specified. | *"I was travelling after the student travel window as I'm an EU student, and the student travel window was very inconvenient. The testing during the travel window was stopped before I needed to get a test in coordination with my travel plans, as the University testing was too early for me so wouldn't have been helpful"* | 5 |
| Inaccessible to key groups | Includes comments about testing facilities being inaccessible to those with additional needs and/or with caring responsibilities. | *"Current testing facilities and practice fail the disabled population"* | 2 |
| Booking issues | Includes comments about students being unable to use the booking system and/or book tests | *Tried to book a slot on website and it was not easy so I gave up* | 5 |
| **Safety concerns** |  |  |  |
| Risk of exposure at the testing site | Comments about concerns of risk of exposure whilst accessing tests | *After watching the virtual tour of the testing facilities (on Instagram), and also showing this to my family, it seemed the booths were all very close together in an enclosed space. This, combined with the high rates of Covid among the student population, made me feel that getting a test in these conditions would put me at greater risk of catching the virus* | 10 |
| Accuracy of tests | Includes comments about tests not being suitable or accurate enough to facilitate safe travel. Also includes comments by students who had had a confirmatory PCR with conflicting result | *The lateral flow tests were advertised as a green card to go home safely without self isolating. It was made to seem like people who test negative are safe. I feel like I was misled because I was not aware that half of positive cases are missed and I felt like I had a false sense of security. Lateral flow tests literally say not for asymptomatic testing on the packaging.* | 11 |
| **Knowledge and understanding** |  |  |  |
| Of testing | Including comments about a lack of /unclear instructions about how to take the test and/or number of tests needed. | *I thought the testing instructions weren't clear enough for someone who isn't familiar with anatomy. "Swab your tonsils for 10 seconds" is only a useful instruction if you know where the tonsils actually are.* | 5 |
| Of eligibility | Includes comments in which participants explain that they did not take part in the testing program as they did not have symptoms/had previously tested positive and/or did not understand who testing was for | *I didn't know the testing facility was for even if you didn't have symptoms.* | 7 |
| **Impact of test results** |  |  |  |
| Lack of support for self isolation | Includes concerns about the lack of support for those who test positive | *"My other main concern is the lack of mental health support for those isolating and/or following all guidelines"* |  |

### Interviews

Twenty students were interviewed; including fourteen who reported that they had taken a test at the university in December 2020 and seven had not. Data is presented under three main themes 1) motives for engaging in testing 2) barriers to testing 3) and using test results to inform behavioural decisions.

#### Motives for engaging in testing

Three main motives for taking part in university testing procedures included 1) to reduce the risk of transmitting the virus 2) for information and 3) following recommendations and guidance.

##### To reduce risk of transmission to others

Most students were more concerned about the risk to others than to themselves (Table 5 quote 1), and were willing to take tests to protect other people from the virus. Tests provided reassurance that they were not spreading the virus to others (quote 2). This was particularly important for those planning to relocate for the holidays (quote 3), those with vulnerable family members (quote 4), or those who considered themselves to have been at risk of exposure to the virus (quote 5).

**Table 5.** Key quotes from interview participants.

| **Motives for engaging in testing** |  |
| --- | --- |
| Reduce the risk of transmission to others |  |
| Quote 1 | "I'm most nervous about passing it on to somebody.... I know a lot of people live with parents or older people or just people on the street. Obviously I don't want to get it myself because that would not be fun but I'm more nervous about passing it onto someone... I'm more worried about hurting someone else" (female, Asian, tested) |
| Quote 2 | "I think it's good for that reason to make sure that you're fine and you know that just going to the shops you're less likely to spread it to someone" (female, Asian, tested) |
| Quote 3 | "Because I was going home, I guess I wanted to lower the chance of me bringing COVID home" (female, white, tested) |
| Quote 4 | "The first time was when the government told us we could all go back home and I wanted to do two tests because if I did get positive and I had to stay here a bit longer, but I would really rather not bring the disease back to my family. Both my parents are a little bit older and my brother's girlfriend is in the vulnerable category" (female, white, tested) |
| Quote 5 | "I had two tests before Christmas because I'm on a PGC programme so I've been in school up until Christmas, then I went to see my family at Christmas" (female, white, tested) |
| For information |  |
| Quote 6 | "I just thought one of the main issues is not knowing whether you have it or not. Information is important so it was an opportunity to get information" (male, white, tested) |
| Quote 7 | "I just wanted to have an idea. I mean I've been pretty good with isolating. I hadn't really been around many people since the beginning of December but... I just wanted to double check, yes... I wanted it for me" (female, mixed ethnicity, tested) |
| Quote 8 | "So I think it's really important just on a mental health level to get tested to make sure that you're not spreading it around. I was negative. I was just worrying for no reason" (female, Asian, tested) |
| Following recommendations |  |
| Quote 9 | "I think just the fact that it was there, so there was obviously the opportunity to [get tested]" (female, white, tested) |
| Quote 10 | "I think I just thought it must be quite important for us to get tested if the University was offering them" (female, white, tested) |
| Quote 11 | "I came back to university and the university asked us all to get tested before our first practical" (female, white, tested) |
| Quote 12 | So in my country, they don't really care about Coronavirus, to put it simply but because my mum is a doctor, she expected me to get tested basically" (female, Asian, tested) |
| **Barriers to the uptake of testing** |  |
| Perceived lack of need |  |
| Quote 13 | "I wasn't getting tested at university because it was people before they were going home. I stayed in [Bristol]" (female, mixed ethnicity, did not get tested) |
| Quote 14 | "I wanted to go home for Christmas so I just isolated to make sure... most of my friends were also isolating and even if they weren't my dad's part of the vulnerable group so it just felt like the proper thing to do" (female, white, did not get tested) |
| Quote 15 | "Most of the others just straight out went to get PCR tests 'cause they were also going back home..." (female, mixed ethnicity, did not get tested) |
| Lack of awareness |  |
| Quote 16 | "To be honest I only became aware of it when I received an email asking me why people weren't – like or asking me why I thought students weren't taking up this offer. So I didn't even know it was there before" (female, mixed ethnicity, did not get tested) |
| Quote 17 | "I don't know if I would have found that information out if I didn't have friends telling me that. I mean I know a lot of people in other places didn't get tested and I don't know if they even knew there was testing going on" (male, white, did not get tested). |
| Access |  |
| Quote 18 | "I couldn't get the links to work and they changed location and something else so it's that sort of booking process and also not knowing where it is that's prevented me from doing it this term" (male, white, did not get tested) |
| Quote 19 | "To be honest, by the time I sort of got round to it and got like, you know, kind of – because you had to get two and one of them I think was clashing with when I was going back [home]" (female, Asian, did not get tested) |
| Risk of exposure at the testing site |  |
| Quote 20 | "What if going to the test centre I come in contact with someone who is positive and I get it there?" (female, white, did not get tested) |
| Quote 21 | "I think practically it was about half an hour walk to the nearest station and because I was already isolating it didn't seem that practical for me to go out and expose myself and then get tested" (female, white, did not get tested) |
| Quote 22 | "I know cases are going up and I'd rather just be in my house where I know I'm safe" (female, white, did not get tested) |
| **Using test results to inform behavioural decisions** |  |
| Quote 23 | "The accuracy of the test is something that I've discussed quite a lot with friends so I was aware that they were not very good at picking up asymptomatic cases, so I feel like I kind of took the negative result with like a pinch of salt" (male, mixed ethnicity, tested) |
| Quote 24 | "I just thought it was like an additional bit of information" (male, white, tested) |
| Quote 25 | "We had this testing I was kind of confident that, well okay I already had those tests. Nobody had any symptoms so I thought, okay it might be okay" (female, Asian, tested) |
| Quote 26 | "I think I was just much less worried about travelling home with COVID. I think I was able to travel home with a bit more sort of like okay, the chances are I probably don't have COVID right now, like I've done everything I can anyway" (male, white, tested) |
| Quote 27 | "I mean I accept that there is a margin for error with anything really but I was willing to accept the results as sufficient, as good enough to make decisions on, like to make my decisions on" (female, white, tested) |
| Quote 28 | "I mean I think it does reassure you doesn't it...it is reassuring because even though it's not very accurate, you still haven't tested positive, so it is a level of reassurance, but it's very, it should be less than what it is, but even though someone who like knows about it and understands, I was still reassured and I think it's hard not to be and I guess isn't that sort of the point of testing anyway" (male, white, tested) |
| Quote 29 | "I definitely wouldn't be visiting anyone who was vulnerable. Everyone in the household I was going to are not in their 60s but I think late 50s max and healthy and young" (female, mixed ethnicity, tested) |
| Quote 30 | Obviously I wouldn't say get tested and go to parties because that's ridiculous but going to the shops and going on a walk and just going to places that you have to be" |
| Quote 31 and 32 | "but then I was very aware that if I went into the supermarket then I could just easily have gone and got infected again so it was like yeah for now but [laugh] 'cause the wording was like at the time you took your test, you tested negative but reinforces like this is very temporary assessment of your situation but it's still better than like having no idea"<br>"my confidence in [the negative test result] decreases with the more contacts I have with people or the more public places I got to or when I'm |
|  | with people. My confidence decreases the more exposure I have to people” (female, Asian, tested) |
| Quote 33 | “I’m sure for some that it would but I’m sure for most that it wouldn’t and I think the people who would probably act differently following one of those negative tests would probably act like that anyway. So I don’t think, for the good impact it would have I think the negative impact would be very small”(female, white, tested) |
| **Recommendations for improving the service** |  |
| Advertising |  |
| Quote 34 | “I think just letting people know about it. Maybe in their [SU] newsletter just mentioning it here and there. I think I only received two emails over the period of a month about it. I think that was a little bit low” (female, Asian, did not get tested) |
| Quote 35 | “To be honest I think it’s quite difficult for the university because a lot of interactions are just dependent on emails because of the virtual situation or the remote situation. I mean I frequently access Blackboard and I do check my emails but sometimes somethings don’t look like they’re relevant to me and so I’ll just like not click or open that. So maybe that’s what happened. Maybe I just thought it was like a generic email, like another COVID generic email been sent out” (female, white, did not get tested) |
| Provision of timely information from trustworthy sources |  |
| Quote 36 | “I read it though I remember thinking it should have flagged up the high false negative rate really, that was one thing throughout the whole thing that I didn’t think was made clear enough. Maybe in the text it should have said even though you tested negative it needs to get across the message that there’s quite a high false negative rate” (female, white, tested) |
| Quote 37 | “The [name of school]specifically did mention the extra, I remember that specifically that they told you a bit more about the test specificity and the Head of the School even offered to have a chat with anyone if they were concerned about it” (female, white, tested) |
| Quote 38 | “The main one being like more information before we go there, I know people that are quite anxious about turning up somewhere, not knowing where they are meant to go or who they are meant to talk to or what they are meant to do, it’s much nicer to have loads of information and then you can pick and choose if you want more information or not then, turning up completely blind and having to try and sort it out” (female, mixed ethnicity, tested) |
| Incentives |  |
| Quote 39 | “I think the important thing is that we have some benefits” (female, white, tested) |
| Quote 40 | “If the Uni sort of sent encouragement saying ‘we want people to get tested’ the I’d do it then”<br>“I know a lot of people who didn’t bother to do the testing just ‘cause they couldn’t be bothered or anything – so there’s definitely some ways that you could influence them, money probably or something like that”(male, white, tested) |
| Quote 41 | “If that could open up me going to the library or going to lectures or anything like that it’s definitely a price worth taking” (female, mixed ethnicity, tested). |

##### For information

In some cases, students wanted to take tests for information (quote 6). Although these students were not necessarily planning to travel, they were keen to take tests for their own benefit (quote 7), including for their mental health (quote 8).

##### Following recommendations

Students reported taking tests simply because they were available (quote 9), and supported by the University (quote 10). For some, tests were a requirement for attendance at in-person lectures (quote 11), or travel (quote 12).

#### Barriers

Barriers to uptake of testing include 1) lack of need 2) lack of awareness 3) access 4) risk of exposure at the testing site.

##### Lack of need

One reason for not engaging in testing was that the student did not think that tests were required or intended for them. For example, one student explained that she had not taken a test at the university because she was not planning to travel away from Bristol (quote 13). Other students were able and willing to isolate, and considered this preferable to testing (quote 14), or demonstrated a preference for PCR tests over LFT (quote 15).

##### Lack of awareness

A lack of awareness prevented some students from accessing the service (quote 16). Students thought that more could be done to promote awareness of testing, particularly among those who do not have a strong network of peers (quote 17).

##### Access

A number of practical barriers were described; including access issues (quote 18), and issues with the timing and location of test sites (quote 19).

##### Risk of exposure at the testing site

Concerns of catching the virus at or on route to the testing centre prevented some students from taking a test (quote 20), particularly among those who had to travel long distances (quote 21). It was noted that cases of the virus were high among the student population, and some considered the risk of exposure to outweigh the benefits of getting tested (quote 22).

#### Using test results to inform behavioural decisions

Most students were very aware of the ongoing debate about the accuracy of LFTs, and reported having discussions with their friends, families, and in some cases, with the university about how accurate the tests were (quote 23). Tests were considered just one piece of information from which to inform decisions (quote 24), often being used alongside other key indicators – such as whether or not the person had been in contact with someone with the virus, of if they had any symptoms (quote 25). Some students reported that testing had reassured them that they had ‘done everything they could’ before travelling (quote 32). Despite limitations, tests were seen as ‘good enough’ to inform decisions (quote 26), and although students reported feeling somewhat reassured by negative test results (quote 29), they described being unlikely to drastically increase contact or to visit anyone considered to be vulnerable (quote 31). Activities were limited to those that were considered essential, such as shopping and exercise (quote 30) and it was recognised that any negative rest result was only “valid” for a limited time, and any subsequent contact was a potential risk (quote 27 and 28).

There was an acknowledgement that receiving a negative test could increase close contact behaviour, but generally it was noted that students who were likely to break the rules would do so regardless of testing status (quote 33).

## DISCUSSION

Our research revealed that one in 10 students had the recommended two LFTs and highlighted demographic disparities in uptake by ethnic group, level of study and year group, and faculty. Data collected from survey and interview participants suggested that whilst students were generally positive about testing, key barriers to uptake remain. Our qualitative data revealed that many participants were motivated to take tests to protect those around them and avoid transmitting the virus to their friends and family. However, students reported a number of barriers to uptake; including a lack of awareness of the testing service, problems accessing the service, a lack of knowledge and understanding of testing procedures, and concerns about the accuracy and safety of testing. Although overall uptake was low, many of those who did not take tests described a lack of need for tests, either because they were not travelling, were unlikely to have been exposed to the virus, were already isolating, or were tested elsewhere.

Mass testing for COVID-19 is relatively new, and results of testing programmes are ongoing. Our data revealed low testing uptake, particularly among those from ethnic minority groups. Similar patterns in testing uptake have been observed with some other public health interventions such as home HIV testing ^20^. The mass COVID-19 LFT pilot conducted in Liverpool also reported a lower test uptake, as well as a higher positivity rate, among those from minority ethnic groups ^21^. The very small number of positive tests during the study period precluded analyses on demographic variations in positivity, both due to a lack of power and the potential for deductive disclosure. Further research is urgently needed to explore barriers to testing among these populations and co-create interventions to support the uptake of tests if and when required.

In line with findings from other universities, students engaging in testing were motivated to do so to protect those around them ^22^. Students were well informed about the limitations of tests, often describing test results as just one piece of information, and using them with caution to inform their behaviour ^23^. Many students had done their own research, had discussions with their friends, family, tutors and lecturers to maximise their knowledge of testing. This highlights the need for improved communications from universities to enable students to make their own informed decisions. Indeed, recent research that has shown basic and simple messages may not be suitable for communicating complex information about how to behave during the pandemic ^24^ and students are likely to appreciate having the opportunity to access information about the sensitivity and specificity of the tests. Despite concerns that testing would increase risky contact, we did not find evidence to support this. Students were well informed about the limitations of the tests and used them with caution to inform behavioural decisions.

A key strength of this research is the use of a mixed methods approach. Additionally, though some other universities have evaluated their LFT programmes ^25, 26^ we are not aware of any reporting data on testing uptake and exploring demographic variations in uptake among the whole student body. This is a unique strength of our work and provides crucial information to inform future university testing strategies. Our work identified several ways in which engagement may be enhanced. As many students had not been aware of the testing service, a persuasive, targeted and personalised advertising campaign may increase uptake. To maximise engagement, advertisements should be co-created with the intended recipients of campaign. Such a campaign should include encouragement from trusted sources, and emphasise the benefits of testing to encourage participation among those who may be apathetic. It would also need to reassure those who are anxious about accessing the testing services.

A limitation of the analyses on testing uptake is that denominator was all students enrolled at the university. The university doesn’t hold comprehensive and reliable information on which students were resident in Bristol during the testing period. However, in our sensitivity analysis in which excluded students who were likely not to be in Bristol at the time of testing the findings were little altered. A key limitation of the survey and interview data is that participant recruitment occurred via social media, and it is likely key communities (e.g., those who do not engage with university managed social media accounts) were missed.

## Conclusions

LFT continues to play an important and expanding role in the UK’s COVID strategy ^3, 4^. If regular LFT is considered appropriate and worthwhile going forwards then work is needed to monitor trends in testing uptake among student, and other, populations. Importantly, we need to strive for equity in access to and uptake of testing. Our findings should be used to inform the wider debate around the usefulness and appropriateness of the widespread use of LFT for asymptomatic people.

## Data Availability

The datasets used and/or analysed during the current study are available from the corresponding author on reasonable request.

## Ethics approval and consent to participate

Ethical approval was provided by the University of Bristol – Ethical approval was obtained from University of Bristol faculty ethics committee (Reference 115084). All interview participants verbally consented to take part in the study.

## Acknowledgements

We would like to thank Beccy Bridges and Dudley Trueman for providing us with University of Bristol student demographics and testing uptake data, and Natalie Seaton-Lucas for providing us with contextual information on the University of Bristol testing programme.

## Supplement 1. Survey

**Demographics**

1. Age [free text]
2. Sex [free text]
3. Ethnic group [free text]

**Living situation**

1. Which best describes you
  - Undergraduate student living on campus
  - Undergraduate student living off campus
  - Postgraduate student living on campus
  - Postgraduate student living off campus
  - Staff
  - Other
2. Student Year group
  - Year one
  - Year two
  - Year three
  - Year four
  - Postgraduate
  - Other /NA
  **Testing**
3. Have you taken a test as part of university testing?
  - Yes
  - No
  **[IF YES TO Q2]**
4. How many tests have you taken as part of university testing?
  - One
  - Two
  - Other
5. How many tests were negative? **Motives**
6. What made you decide to get tested? (tick all that apply)
  - I want to return to “normal”
  - I want to protect others
  - I want to protect myself
  - I want to see vulnerable family members
  - My friends and/or family asked me to get tested
  - It gives me peace of mind
  - People I know are doing it
  - My household were doing it
  - I have symptoms
  - I am curious
  - I want to help to fight the virus
  - I think it’s the right thing to do
  - I was told to get tested
  - The testing site is convenient
  - Other
  **Testing experience**
7. How easy was it to be tested?
  - Very easy
  - Somewhat easy
  - Neither easy nor difficult
  - Somewhat difficult
  - Very difficult
8. If difficult, please describe what the difficulty was (free text) **Attitudes to testing**
9. What would you say your view of repeat testing is?
  - Very negative
  - Somewhat negative
  - Somewhat positive
  - Very positive
10. How likely are you to get tested again?
  - Very unlikely
  - Unlikely
  - Neither likely or unlikely
  - Likely
  - Very likely
11. What might stop you being tested in the future?
  - If I don’t have time
  - If my allocated slot isn’t convenient
  - I’m worried I might have to wait a long time at the testing site
  - The location of the test was not convenient
  - The testing experience was unpleasant
  - I’m worried about catching the virus by going for a test
  - I’m worried about having to self-isolate
  - I’m worried I wouldn’t have enough practical support if I needed to self-isolate after a positive test
  - I don’t know how this will help me or the people around me
  - I’m worried that this is a waste of resources
  - I don’t think it’s important
  - I think this might take away tests from someone who needs it more than me
  - I don’t think the tests are accurate
  - The people I live with don’t want me to get tested
  - I’m worried about the impact on my household if I test positive
  - I’m worried I might have to provide details of my contacts if I test positive
  - I’m concerned about who will have access to my data
  - I am concerned about how my data will be used
  - Other, please state
12. If you tested positive, how easy would it be for you to self-isolate?
  - Very difficult
  - Difficult
  - Neither easy nor difficult
  - Easy
  - Very easy
  **Impact of test result**
13. Which of these statements best describes what a negative test means
  - The person is definitely not infectious
  - The person is probably not infectious
  - The person is probably infectious
  - The person is definitely infectious
  - I don’t know
14. Thinking about the 14 days after your <u>last</u> test result, how often have you followed government advice on social distancing?
  - All of the time
  - Most of the time
  - Some of the time
  - Not at all

## Supplement 2 – semi-structured topic guide

[Students who did engage in the testing service offered by the University]

1. What made you decide to get tested the university?
  - What was your initial reaction to university testing?
  - Did you have any concerns about getting tested?
  - Were any of your household planning to have a test?
  - How important / necessary?
2. Can you tell me about your experience of getting tested?
  - What was it like?
  - What worked well?
  - What was the most difficult part of testing?
3. What did you think of the messaging you received about testing?
  - What information did you receive about how to take the test?
  - What information did you receive about the test result?
  - Was anything unclear or confusing?
  - What (if anything) was missing?
4. How did you feel when you received a negative test result?
  - What did this mean for you?
  - What did this allow you to do?
  - What didn’t this allow you to do?
  - Did you have any concerns about your result?
5. Worried about testing positive?
6. How problematic would a positive test result have been?
7. If necessary, would you be willing to get tested again?
  - Why?
  - What might influence this decision?
  - Are there any situations that it would be particularly important to get tested?
  - What could be done to make it better / easier for people to test/isolate?
8. What is it like being a student at this time?
9. Is there anything else you would like to say about asymptomatic testing? [for those who tested positive]
  - What did you do when you received a positive test result?
  - What did this mean to you?
  - What does the term self-isolation mean to you?
  - Can you tell me about your experiences of having to self-isolate?
  - What steps did you take?
  - What was the most difficult part of having to self-isolate?
  - What did you do to overcome any problems you had?
  - What would have helped you overcome any problems that you had?
  - Do you think having to self-isolate had any impact on your health/wellbeing in anyway?
  - Were there any times that you were not able to self-isolate?
  - Why? What happened?
  - What support did you have to help you with daily testing and self-isolation?
  - What did you think of the support?
  - What support did you need? / what was missing?
  [Students who did not engage in the testing service offered by the University]

1. What made you decide not to get tested the university?
  a. What was your initial reaction to university testing?
  b. Did you have any concerns about getting tested?
  c. How important / necessary do you think it is to be tested?
  d. Were any of your household planning to have a test?
2. How problematic would a positive test result have been?
3. What (if anything) would make you more likely to be tested in the future?
  - Practical / logistical support
  - Reassurance
  - Social pressure
  - Incentives
4. What did you think of the information or advice you had about testing?
  - How clear was the information about how to take the test?
  - How clear was the information about the test result?
  - Was anything unclear or confusing?
  - What (if anything) was missing?
5. If necessary, would you be willing to get tested again?
  a. Why?
  b. What might influence this decision?
  c. What could be done to make it better / easier for people to test/isolate?
6. Is there anything else you would like to say about asymptomatic testing?

## Supplement 3. Demographic data by testing group among survey respondents

| Characteristic | Participated in testing N = 328 | Did not participate in testing<br>N = 108 |
| --- | --- | --- |
| Undergraduate student on campus | 187 (57%) | 38 (35%) |
| Undergraduate student off campus | 96 (29%) | 38 (35%) |
| Postgraduate student on campus | 14 (4%) | 15 (14%) |
| Postgraduate student off campus | 30 (9%) | 15 (14%) |
| Female | 222 (68%) | 67 (62%) |
| Male | 77 (23%) | 17 (16%) |
| Prefer to self describe | 3 (1%) | 2 (2%) |
| Gender not provided | 26 (8%) | 22 (20%) |
| Ethnic minority group | 38 (12%) | 35 (32%) |
| White | 267 (81%) | 56 (52%) |
| Missing | 23 (7%) | 17 (28%) |
| Total | 328 | 108 |

